# A Mixed Methods Comparison of Artificial Intelligence-Powered Clinical Decision Support System Interfaces for Multiple Criteria Decision Making in Antidepressant Selection

**DOI:** 10.1101/2022.10.03.22280635

**Authors:** Akiva Kleinerman, David Benrimoh, Grace Golden, Myriam Tanguay-Sela, Howard C. Margolese, Ariel Rosenfeld

## Abstract

**BACKGROUND:** Artificial intelligence-powered clinical decision support systems (AI-CDSS) have recently become foci of research. When clinicians face decisions about treatment selection, they must contemplate multiple criteria simultaneously. The relative importance of these criteria often depends on the clinical scenario, as well as clinician and patient preferences. It remains unclear how AI-CDSS can optimally assist clinicians in making these complex decisions. In this work we explore clinician reactions to different presentations of AI results in the context of multiple criteria decision-making during treatment selection for major depressive disorder.

**METHODS:** We developed an online platform for depression treatment selection to test three interfaces. In the probabilities alone (PA) interface, we presented probabilities of remission and three common side effects for five antidepressants. In the clinician-determined weights (CDW) interface, participants assigned weights to each of the outcomes and obtained a score for each treatment. In the expert-derived weights interface (EDW), outcomes were weighted based on expert opinion. Each participant completed three clinical scenarios, and each scenario was randomly paired with one interface. We collected participants’ impressions of the interfaces via questionnaires and written and verbal feedback.

**RESULTS:** Twenty-two physicians completed the study. Participants felt that the CDW interface was most clinically useful (H=10.29, p<0.01) and more frequently reported that it had an impact on their decision making (PA: in 55.5% of experienced scenarios, CDW: in 59.1%, EDW: in 36.6%). Clinicians most often chose a treatment different from their original choice after reading the clinical scenario in the CDW interface (PA: 26.3%, CDW: 33.3%, EDW: 15.8%).

**CONCLUSION:** Clinicians found a decision support interface where they could set the weights for different potential outcomes most useful for multi-criteria decision making. Allowing clinicians to weigh outcomes based on their expertise and the clinical scenario may be a key feature of a future clinically useful multi-criteria AI-CDSS.

## 1. Introduction

Clinical decision support systems (CDSS) are health information technologies that provide data and recommendations to assist clinicians in medical decision making (Sutton et al., 2020; Zikos and DeLellis, 2018). Recent improvements in artificial intelligence (AI) have resulted in the proliferation of AI-powered CDSS (AI-CDSS) with the capability to leverage data and automate observations to produce novel predictions (Sutton et al., 2020).

One of the many fields to which AI-CDSS are currently being applied is the treatment of major depressive disorder (MDD) (Benrimoh et al., 2018; Kleinerman et al., 2021; Mehltretter et al., 2020, 2019; Tanguay-Sela et al., 2022) Depression is highly prevalent in the general population and is associated with significant morbidity and mortality (Lépine and Briley, 2011). Despite its high prevalence and the availability of many effective treatment modalities, only a third of patients will reach remission after their first treatment (Warden et al., 2007). Antidepressants are frequently prescribed using an educated trial-and-error approach (Tomlinson et al., 2020) in which patients typically undergo several rounds of different antidepressants before achieving remission, and there are no current widely-used tools which can improve the match between patients and antidepressants at the point of care without expensive additional testing (Lépine and Briley, 2011; Tanguay-Sela et al., 2022).

Medical decisions, such as which antidepressant should be chosen for a given patient, are complex and require the consideration of many possible outcomes, such as treatment response or side effects, which can be difficult to predict. In an effort to improve decision making, researchers have developed artificial intelligence models that predict treatment-specific probabilities of remission for individual patients (Kleinerman et al., 2021; Mehltretter et al., 2020, 2019). These tools were integrated into CDSS and found to have the potential to improve treatment outcomes (Benrimoh et al., 2021; Lépine and Briley, 2011; Tanguay-Sela et al., 2022). In addition, recent work has shown potential in predicting the occurrence of side effects in response to various treatments based on genetic, clinical, and demographic data (Busch and Menke, 2019; Taliaz et al., 2021).

Even if models are generated which can accurately and reliably predict side effects, remission probabilities, or other outcomes, an important question would remain: how should this information be presented to clinicians to best support decision making? This is not a trivial question; if information is not presented to clinicians in a manner that is judged by them to be useful, trustworthy, and easy to understand and explain to patients, the likelihood that they will actually use this information in clinical decision making drops significantly (Bussone et al., 2015; Murphy, 2014; Sim et al., 2001). We argue that the fundamental issue to consider here is as follows: when choosing a treatment, a clinician must determine, usually together with the patient, the importance of the various outcomes and criteria and aggregate all this information to produce a (usually implicit) ranking of the possible treatments accordingly. In addition, there are usually several treatment options, and preferences and existing biases of patients and clinicians vary. Given this complexity, we consider the question of what format the outputs of clinically useful machine learning predictions should take in a multi-criteria context, whether these should be raw probabilities or summary scores, and, in the case of summary scores, what kind of weighting process clinicians would most appreciate to reflect the importance of each criterion in the decision.

In order to inform our approach to this question, we turn to the field of multiple-criteria decision making (MCDM), a field of research that addresses decision making that includes multiple, often conflicting, criteria (Aruldoss et al., 2013). Note that ‘criteria’ in the healthcare context often refers to outcomes, such as side effects or treatment response. Work in this field has proposed and evaluated various methods for aiding individuals or groups of decision makers. Some of these methods explicitly combine criteria in a way that reflects the decision makers’ preferences (Zavadskas and Turskis, 2011). Similar to many MCDM applications, healthcare decisions often involve multiple conflicting criteria, and the stakes are naturally high. Recently MCDM methods have been applied extensively within healthcare (Baltussen et al., 2019; Thokala et al., 2016) and guidelines have been established to best apply MCDM methods in healthcare decisions (Thokala et al., 2016). Previous research has concluded that there is no optimal MCDM method that best fits all healthcare decisions, and that the choice of MCDM method should depend on the specific context of the decision and the objectives of care (Baltussen et al., 2019; Marsh et al., 2017; Thokala et al., 2016).

Prior work has evaluated CDSS and AI-CDSS that provide information about different criteria, such as side effects or remission probabilities (Popescu et al., 2021; Qassim et al., 2022; Trivedi et al., 2004), and some work has implemented CDSS which have a weighting of criteria based on expert opinion (Chen et al., 2012). However, to our knowledge, no previous work has investigated how best to present the results of an AI-CDSS aimed at assisting in MCDM by comparing several different methods of presenting this information to clinicians.

## 2. Methods

We tested three AI-CDSS interfaces focused on supporting clinicians when making the multi-criteria decision of choosing an antidepressant for a patient with a MDD. Our objective was to determine which type of data presentation clinicians find most helpful. We developed three prototype interfaces. Each interface was randomly paired with a clinical scenario of a depressed patient as well as probabilities of remission and three common side effects for five commonly used antidepressants. The interfaces differed in the way they supported the weighing of the various criteria. The first, “probabilities-alone” interface provided only the probability for each outcome separately. The second, “clinician-driven” interface allowed clinicians to determine patient-specific weights for all four criteria (i.e., remission and three side effects) and generate a summary score for each antidepressant, which aggregates the criteria based on the clinician’s weights into a single score for the treatment (further described below). The third “expert-driven” interface presented the same weighting procedure and ranked summary score, except with weights pre-selected based on expert opinion (see below). We hypothesized that clinicians would find the “clinician-driven” interface most useful because it provided the utility of the summary score while also allowing them to exercise their clinical judgment. In addition, this interface has the most flexibility and is most adaptable to each individual patient, raising the possibility that it might, of the three interfaces, be most useful for clinicians in the context of shared decision making (Elwyn et al., 2012) with their patient.

### 2.1 Participants and Recruitment

Participants were psychiatrists and primary care physicians (PCPs) from Israel, Canada and the USA. Twenty-eight participants (17 psychiatrists and 11 PCPs) were recruited via email and LinkedIn^1^.

We recruited participants for this online experiment between June and August 2022. Most participants completed the experiment on their own, though all participants were offered to schedule a video conference with an investigator to assist them in case they had any questions. Three participants elected to participate in video conferences. The study was approved by the Committee for the Approval of Research Involving the Participation of Human Subjects at Bar-Ilan University. Participants provided written informed consent and, if they chose, were entered into a draw to win one of three gift cards as compensation for their time. Participants who participated through video conferences were compensated for their time but were not included in the draw.

### 2.2 Study Design

The study flow is described in Figure 1. Participants first provided informed consent, then read a brief introduction to the experiment and completed a demographic questionnaire. Participants then completed three phases in which they were asked to make a treatment decision, each phase included a clinical scenario and a clinical decision support interface. Participants filled out a questionnaire after each of these phases which asked them about their treatment choices during the phase, their understanding of the interface they used, and the impact of the interface on their choice. After the three phases, they completed an exit questionnaire that included general questions about the experiment as well as questions about specific interfaces (Supplementary Material Section 7).

**Figure 1:**
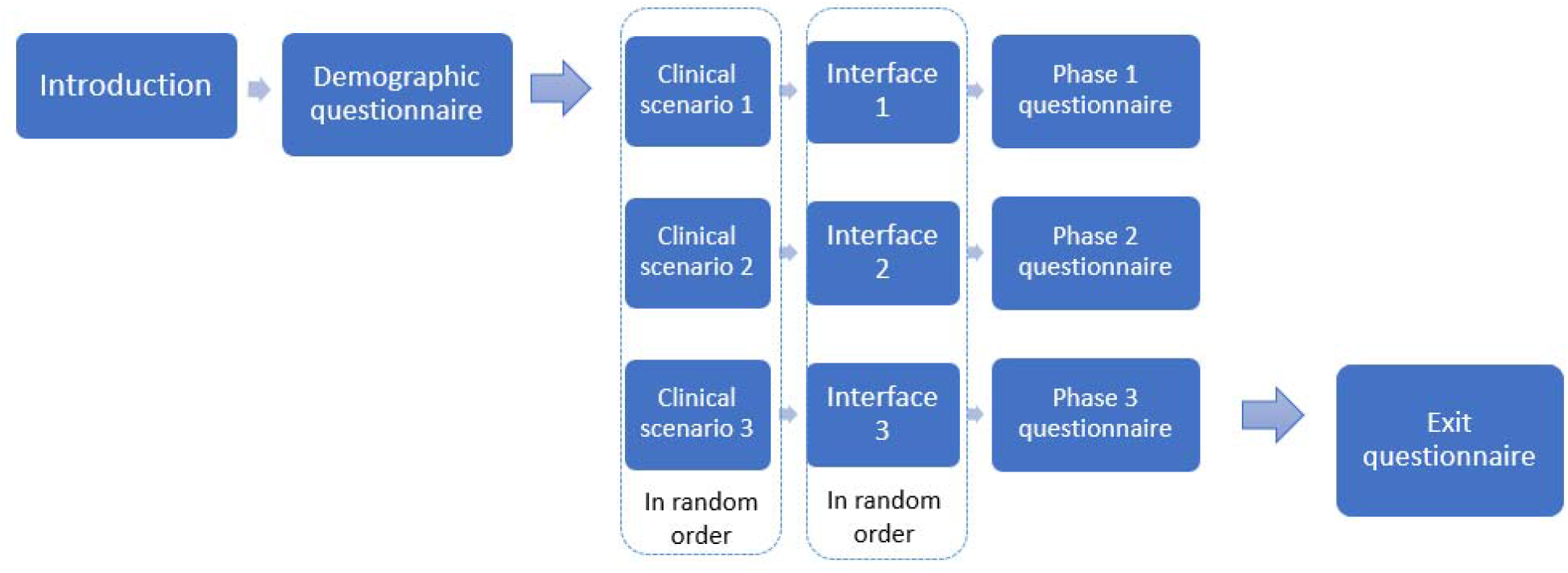
The experiment process. The experiment included three phases, each phase consisted of a clinical scenario and an interface for assisting treatment selection.

### 2.3 Materials

We created a simulated clinical decision support tool, with which clinicians interacted in each phase. The tool is a web-based platform developed using Anvil^2^ and can be accessed at: *https://4esgtei3kqpigzig.anvil.app/Z7DDNV6PSZ77TIRIYUX2FUKM*. This was a simulated tool aimed at testing different interface types. This platform included three decision support interfaces as well as three clinical scenarios, further described in the Supplementary Material (Section 2). Participants experienced all three clinical scenarios in random order, and each was randomly paired with one of the three interfaces.

#### 2.3.1 The interfaces

The study included three interfaces for assisting with the treatment decision. Before each interface, the user was presented with a written clinical scenario and estimated probabilities for remission and for occurrence of three side effects (weight gain, sexual dysfunction, and fatigue) for five antidepressants (see Supplementary Material Section 2.3). The probabilities were presented visually with an interactive bar chart that compared the various outcomes (see Figure 2).

**Figure 2:**
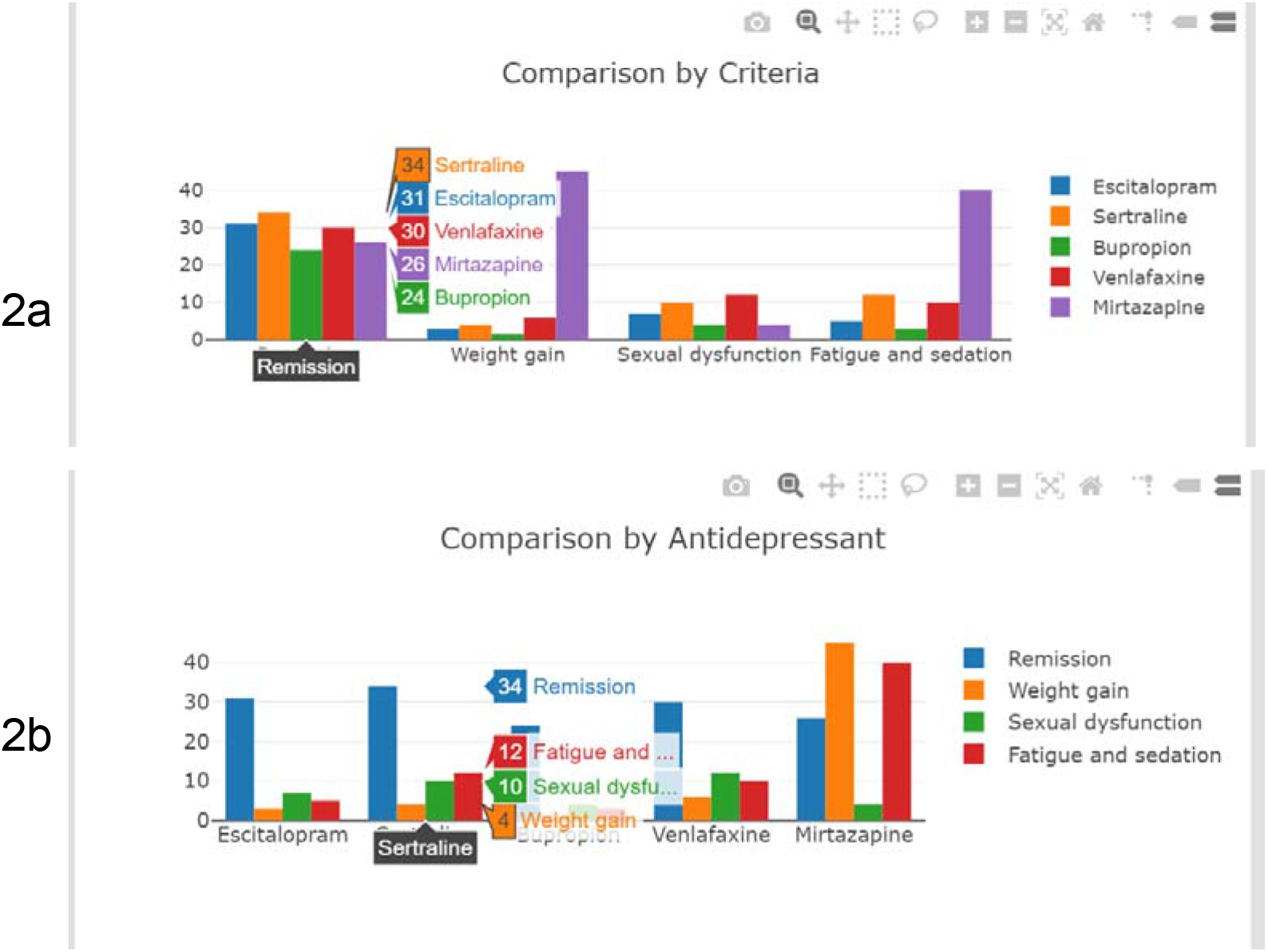
Interactive visualization of predictions. Hovering over the bars reveals more detailed information about the predictions. A drop-down box at the top of the chart enables switching between two views: in the first (2a), the bars are grouped by outcome and in the second the bars are grouped by medication (2b).

In the “probabilities alone” (PA) interface, no additional information was presented beyond the patient’s clinical data and probabilities depicted in a bar graph and in table form. In the “clinician-determined weights” (CDW) interface, users were presented with an interactive tool for assigning importance-weights to each of the outcomes, ranging from 0 to 10 (see Figure 3). After setting weights, they were able to generate a recommendation in the form of a list of medications ranked by a summary score that was based on their weighting. In addition to the ranked list, there was a stacked bar graph that presented the summary scores and the various components that formed the scores (see Figure 4). Finally, in the “expert-determined weights” (EDW) interface, users were presented with a weighting of the outcomes that was derived from experts’ weighting of the scenario. After that, the interface presented a recommendation based on the experts’ weighting - the same kind of ranked list and stacked bar graph as in CDW. The weights in this interface were determined by three psychiatrists with experience in depression treatment. Note that to make the EDW interface more realistic, the participants were told that the weights were derived from weighting of *similar* patients, and not the patients themselves.

**Figure 3:**
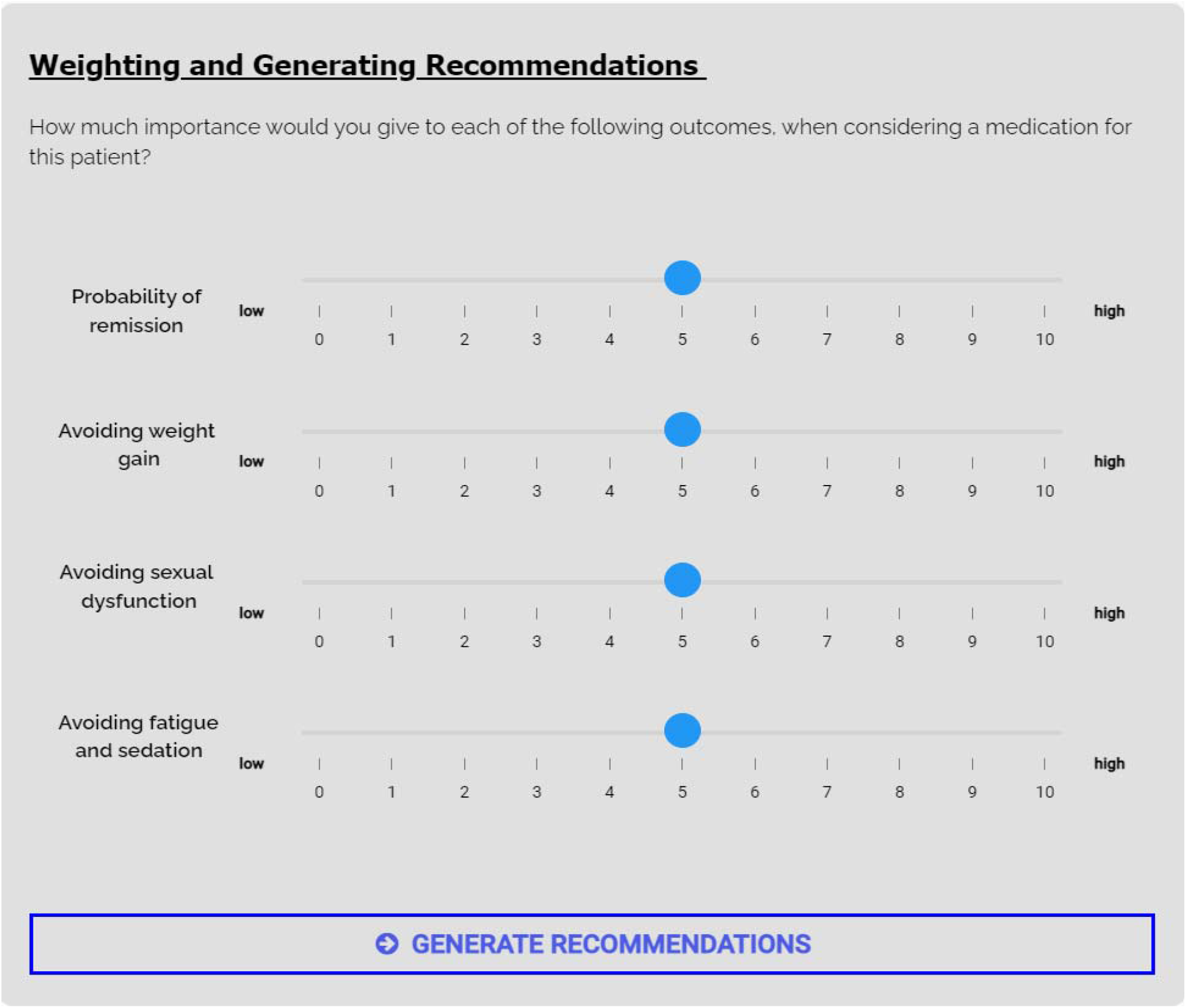
setting weights in the CDW interface. The user set a weight for each outcome using the scales.

**Figure 4:**
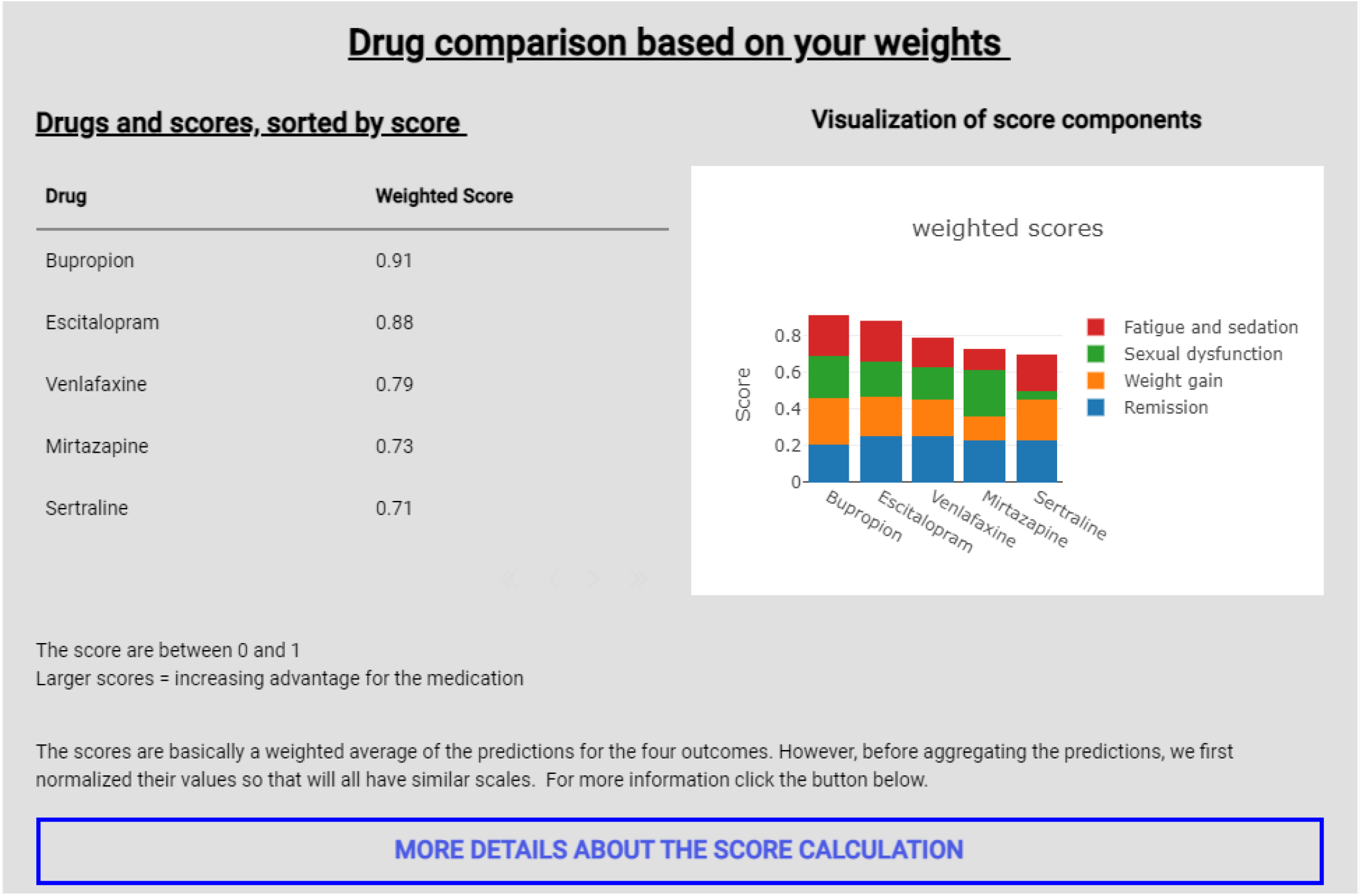
Drug ranking in the CDW interface. The drugs are sorted according to the user’s weighting.

The weighted summary score calculation was based on the weighted sum model (WSM) method (Thokala et al., 2016; Triantaphyllou, 2000), a widely used and intuitive method for MCDM. The users were given the option to read more about the scoring method; this additional information is presented in the Supplementary material (Section 8).

Participants were asked to choose a medication for the patient before moving on to the next phase. They were also asked what medication, if any, they had intended to select after reading the clinical scenario but before reviewing the decision support elements of the interface.

#### 2.3.2 Side effects and antidepressants

The side effects that were considered in the experiment were weight gain, sexual dysfunction and fatigue. We chose these side effects as they are commonly identified by patients as reasons for non-adherence (Ashton et al., 2005; Burra et al., 2007; Serretti et al., 2010) and are often the subject of discussion between patients and clinicians when selecting treatment. Including more side effects would likely have made it more practically difficult for clinicians to process the information given the novelty of the interface. The medications presented were escitalopram, sertraline, bupropion, venlafaxine, and mirtazapine. These are representative of commonly used antidepressant classes, have been well-established in clinical use for several years, and are considered to be first-line antidepressants (Kennedy et al., 2016).

### 2.4 Analysis

#### 2.4.1 Quantitative analysis

Our primary question of interest was to determine which interface participants would judge to have the greatest clinical utility and which would have the most impact on treatment decisions. We evaluated this based on the exit questionnaire and the three phase questionnaires. These were all custom made, given the novelty of this study. The phase questionnaires were intended to capture participants’ immediate impressions after using each interface, while the exit questionnaire assessed their overall experience using the platform after having gone through the three phases, and more specifically their thoughts on its perceived clinical usefulness. The questionnaires are presented in the Supplementary Material (Section 7).

Due to the relatively small sample size, the values were not expected to distribute normally. Therefore, statistical analysis was performed with the Kruskal– Wallis test (McKight and Najab, 2010), which does not assume normality, and Dunn’s test (Dunn, 1964) was used for post-hoc pairwise comparison. This study was exploratory and as such did not include corrections for multiple comparisons.

We first examined the differences between the phases in the whole sample, and then divided the sample by physician specialty to interrogate differences between psychiatrists and PCPs.

Further analyses examining trust, perceived understanding, impact of the clinical scenarios on treatment choice, desire to use a decision support system in general, time spent on the different interfaces, and further between-group differences are presented in the Supplementary Material.

#### 2.4.2 Qualitative analysis

We performed an exploratory qualitative analysis to supplement the quantitative analysis. The qualitative data consisted of the written feedback from the participants in the questionnaires (see Supplementary Material Section 7) and the transcribed recorded comments from the participants who took part in the video conferences. Note that providing written feedback in the questionnaires was not mandatory for completing the experiment. We used inductive thematic analysis (Guest et al., 2011) for the qualitative analysis process. We initially identified eight themes: clinical utility, impact on decision making, weighting of criteria, concerns and difficulties, ease of use, interface features, shared decision making and experiment design. Each theme had several related sub-themes. We employed the investigator triangulation method (Archibald, 2016) during the qualitative analysis, in which two researchers individually coded the qualitative data and later compared the codes. The researchers independently read and coded fragments of the qualitative data into sub-themes. Redundancies were eliminated by merging some of the themes and rearranging the sub-themes. The coders then independently reread and coded all the data into a final summary table.

## 3. Results

### 3.1 Quantitative Results

#### 3.1.1 Sample description

Our sample consisted of 28 participants who consented, and 22 who completed the experiment. Within the 22 participants there were 13 psychiatrists and 9 PCPs. Table 1 presents the demographics of the participants who completed the experiment.

**Table 1:** Demographic statistics: for the participants who completed the study.

| Attribute | Measure | Value |
| --- | --- | --- |
| Age | N | 22 |
|  | Mean | 47.3 |
|  | Std | 8.53 |
| Gender | female | 11 |
|  | Male | 11 |
| Years of experience | 0-4 | 2 |
|  | 5-9 | 6 |
|  | 10-14 | 2 |
|  | 15+ | 12 |
| Specialty | Psychiatry | 13 |
|  | Family Medicine/<br>Primary Care | 9 |
| Training location | Israel | 10 |
|  | USA | 6 |
|  | Canada | 4 |
|  | Other | 2 |
| Practice environment | Outpatient clinic | 11 |
|  | Hospital outpatient department | 5 |
|  | Inpatient service | 3 |
|  | Specialized mood disorder service | 1 |
|  | Other | 2 |

#### 3.1.2 Clinical utility of the interfaces

Participants were asked how clinically useful they found each of the interfaces using a 5-point Likert scale (1= very little, 5= very much). Using the Kruskal-Wallis test, we found a significant difference between the interfaces (H=10.29, p<0.01). Using Dunn’s test, we found that the CDW interface (mean: 4.18, SD: 0.89) outperformed both the PA (mean: 3.27, SD: 0.86) (p<0.01) and EDW interfaces (mean: 3.41 SD: 1.19; p<0.05). No other significant differences were found across the phases. Similar results were also found when grouping the sample into psychiatrists and PCPs (see Supplementary Material Section 3).

#### 3.1.3 Perceived Impact of the Interface on Decision Making

In each phase participants were asked if the specific interface they used had an impact on their treatment decision and answered “Yes” or “No”. We found that after using the PA interface, 54.5% of the participants answered that the interface impacted their decision. However, there was a notable difference based on specialty: only 38.5% of psychiatrists reported an impact of the PA interface on their decision compared to 77.8% of PCPs. After using the CDW interface, 59.1% of participants reported an impact on their decision-making; this was true for 53.8% of the psychiatrists and 66.7% of the PCPs. Finally, only 36.4% of the participants reported an impact of the EDW on their decision-making; 23.1% of the psychiatrists said the EDW interface impacted their decision-making, compared to 55.6% of the PCPs. All differences did not reach statistical significance, likely due to the small sample size. Table 2 present these results.

**Table 2:** **Percentage of participants who reported that the interface impacted their drug choice** in the full sample and in the psychiatrist and PCP groups. Overall, the impact on the treatment decision was smaller in the psychiatrist group.

| Interface | All participants | Psychiatrists | PCPs |
| --- | --- | --- | --- |
| Probabilities Alone (PA) | 54.5% | 38.5% | 77.8% |
| clinician-determined weights (CDW) | 59.1% | 53.8% | 66.7% |
| expert-derived weights (EDW) | 36.4% | 23.1% | 55.6% |

#### 3.1.4 Change in Treatment Selection

Next, we analyzed participants’ change in treatment choice in the various interfaces. Overall, in 84.8% of the scenarios, the participants reported that they had an initial choice before viewing the interface. We found that 25% of the initial decisions changed after viewing the interface. In the PA interface, 26.3% of the participants (who had an initial choice) changed their choice, 33.3% in the CDW interface and 15.8% in the EDW. PCPs tended to change their initial choice more often than psychiatrists: overall, across all interfaces, 30% of PCPs changed their initial choice, compared to 22% of psychiatrists. Further details can be found in the Supplementary Material (Section 4).

### 3.2 Qualitative Results

Sixteen participants provided qualitative data. Of these, three participated through video conference and provided additional verbal comments. We reduced the original eight themes to the following five: 1) clinical utility; 2) weighting of criteria; 3) impact on decision-making; 4) ease of use and concerns; and 5) experiment design. All themes had several sub-themes. Given our research interest, we discuss the first three themes below; the additional themes are discussed in the Supplementary Material.

#### 3.2.1 Clinical Utility

Nine participants referred to the clinical utility of the various interfaces. Five of the participants expressed willingness to use such a system in their own practice and two participants mentioned that they would not use the system themselves, but that they believe it could be useful for other healthcare providers. One psychiatrist commented: “I think giving expert weighed decisions could be very helpful for PCPs or doctors who may not have specialized much in psychiatric training”. One participant suggested that nurse practitioners could use the system. Two participants mentioned that the CDW interface could support shared decision-making. To this effect, one participant commented: “I think it would be particularly useful to use it as a shared decision-making tool. I think that the patients would love to see how their doctor comes to their decision about medication choice.” Two participants said they would only use the part of the interface that presented the probabilities for the various criteria and would not use the additional weighted summary score component. As one physician mentioned: “I would use it for the raw probabilities, as I would weigh things myself based on my experience “. Three participants reported that the presentation of the probabilities for the side effects was helpful. For example, one participant noted: “It was important for me to know about fatigue - I didn’t know exactly the percentages of side effects and the data helped me decide”.

#### 3.2.2 Weighting of Criteria

Eight participants commented specifically about the criteria weighting, which was a component of CDW and EDW interfaces.

##### Clinician weighting

Overall, the participants had a positive attitude towards performing criteria weighting: five of eight participants reported that the process of explicitly setting weights for the criteria in the CDW interface helped them to compare the treatments, and to make sure they did not overlook or undervalue any specific criterion. One participant commented: “it made me consider the overall fit of the medication I chose, rather than focusing on one aspect — difficulty falling asleep — that might not have fit very well for this patient.” One participant wondered if it would be possible to “add an option for more detailed weighting for specific dimensions of remission”. On the other hand, one participant commented that the weighting process is too simplified in the CDW interface: “Weighting is a complex issue, you also have to take into account the temporality of the side effects and the ability to treat them.”

##### Expert weighting

Three participants expressed discomfort with the weighting component of the EDW interface. One participant said that weighting outcomes is “precisely the clinical work that psychiatrists should be doing”. Another participant disagreed with a specific weight given by the experts. Two participants felt that the EDW interface contradicts the shared decision-making approach, as one clinician wrote “It is clear from previous research that ‘expert’-set weights are no substitute for engaging patients in their own care, shared decision-making, etc”. Notably, all the participants who had a negative comment about the EDW were all psychiatrists. Having said that, one psychiatrist commented that they preferred the EDW over the CDW because “What was the point of the AI if I set the weights myself?” and that the EDW felt like “magic”.

#### 3.2.3 Impact on treatment decision

Thirteen of the participants reported that the interface had an impact on their decision process. Six participants reported that at least one interface confirmed their initial decision. One of these participants reported: “It confirmed the decision that I had in mind… made me more confident that I was not overlooking or under- or over-weighting important information”. Five participants reported that an interface made them change their mind. For example, one participant mentioned: “I changed the initial treatment based on the side effects”. Two participants mentioned that although the interface did not make them change their mind, it made them reconsider their decision. Another two participants said that the interface provided them with an idea for a second-line treatment if needed.

## 4. Discussion

This study aimed to determine which kind of AI-CDSS interface clinicians found most useful in order to provide a base of evidence for further development. We found that, overall, clinicians felt the CDW interface was more clinically useful than the other interfaces. The fact that the clinicians preferred the CDW over the PA interface implies that clinicians appreciate support in aggregating predictions about multiple criteria into actionable summary scores. The clear preference of CDW over EDW can potentially be explained by two key factors, supported by our qualitative findings. First, the CDW interface allowed for greater clinician autonomy, and this has been found to influence CDSS acceptance (Khairat et al., 2018). Second, the CDW interface was perceived as being more supportive of shared decision-making (Elwyn et al., 2012) as this interface could be used during an exchange with a patient to collaboratively set weights, which is not possible in EDW.

We also found that the CDW interface was most frequently perceived as having an impact on decision-making, very closely followed by the PA interface. This suggests that interfaces which allow for clinicians to exercise their decision-making in terms of which criteria they consider most important are judged to be more impactful than those which apply weighting based on expert opinion, and that there may be an advantage in this respect for an interface which provides a summary score, though this advantage may be small. Similarly, we found that the CDW interface was most likely to change participants’ initial decisions. These differences were not significant, likely because of the small sample size. Some participants commented that the CDW interface did not change their decision, but rather confirmed their initial decision and helped them make sure they were thoughtfully considering all criteria. In addition, as might be expected, we found that psychiatrists were overall less likely to report an impact of the interfaces on their decision in comparison to PCPs, with this discrepancy being most pronounced in the EDW. These results suggest that the interfaces may have the greatest clinical utility for primary care workers, in line with other AI-CDSS research (Benrimoh et al., 2021; Tanguay-Sela et al., 2022), though there may still be utility for psychiatrists as long as the CDSS allows them to exercise their expertise in the weighting of criteria.

### Limitations

An important limitation of our work is the small sample size which limited power to detect significant differences in the whole sample and between specialty groups. In addition, we did not correct for multiple comparisons. As such, larger studies are needed to confirm these results. In the qualitative analysis, some participants did not provide any comments and many of the comments were brief. Additionally, the qualitative data may be biased towards the perspective of the participants who participated in a video conference, since they naturally provided more comments. Another limitation is that the platform only considers a subset of the criteria that may affect treatment selection- for example, it only considers three possible side effects, and does not consider factors such as medication cost or medical comorbidities and drug-drug interactions. Additionally, the platform only presented five antidepressants when over a dozen first line treatments are available, and it does not consider treatment choices later in a patient’s evolution, such as augmentation strategies. Finally, this experiment did not include patient contact, which may have changed clinician perceptions of the platform because of their interaction.

## 5. Conclusion and future directions

In this study, we evaluated various interfaces for supporting clinicians while selecting MDD treatment, a complex and multi-criteria decision. To the best of our knowledge, this is the first work that investigated AI-CDSS for MDD aimed at comparing the clinical utility of multiple methods of presenting AI-CDSS data. We discovered that clinicians, and especially PCPs, found decision support interfaces useful for MCDM, and that they were most accepting of an interface that enables them to set weights for various outcomes and to receive treatment recommendations accordingly. In addition, this interface seemed to have the most impact on treatment decisions. Future work should determine the impact of these kinds of interfaces on shared decision-making with real or simulated patients. In addition, future work should compare various methods for MCDM beyond the simple weighting algorithm used here.

## Data Availability

All data produced in the present study are available upon reasonable request to the authors

## 6. Authors’ Contributions

AK and DB contributed equally to this paper. AK conceptualized the experiment, created the experiment platform, conducted the analysis, and wrote the manuscript. DB helped conceptualize the experiment and the platform, helped conceptualize the analyses, helped write the manuscript, and provided supervision. GG helped perform the qualitative analysis and helped write the manuscript. MTS helped to write and edit the manuscript. HM helped to validate the experimental platform and provided guidance on the experiment and reviewed the manuscript. AR helped conceptualize the experiment, reviewed the manuscript, and provided supervision.

## 7. Acknowledgements

## Data availability

All data required to replicate the experiment is provided in the manuscript and supplementary material; data collected for this experiment is available on request for non-commercial use.

## Summary Table

|  |  |
| --- | --- |
| **What was already known on the topic** | • Clinical decision support systems (CDSS) have increasingly been the subject of research in a number of disorders, such as major depressive disorder (MDD). Previous research has shown that integrating artificial intelligence models into CDSS (AI-CDSS) has the potential to improve treatment outcomes and processes in conditions such as MDD.<br>• Making treatment decisions, such as selecting an antidepressant, requires clinicians to weigh risks and benefits related to multiple outcomes, such as symptom improvement and side effects.<br>• Most work on AI-CDSS has focused on the provision of probabilities about single outcomes, such as the probability of remission, but do not provide clinicians with support in integrating probabilities for different outcomes in order to reach a decision.<br>• While it is clear from previous studies that many clinicians appreciate having access to an AI-CDSS, the optimal manner of presenting data and predictions to them in a multi-criteria decision-making process remains unclear. |
| **What this study added to our knowledge** | • In this study, we have demonstrated that clinicians can use interfaces to weight different possible outcomes to facilitate multi-criteria decision-making<br>• We also now know that clinicians prefer to set weights or review raw predictions themselves, rather than accepting expert recommendations for weighting<br>• Clinicians may view systems that allow them to weight criteria in order to arrive at a decision as a vehicle for shared decision-making, a hypothesis which must be tested in future research |

## Footnotes

1 http://linkedin.com

2 https://anvil.works/

